# Evaluation of an emergency safe supply drugs and managed alcohol program in COVID-19 isolation hotel shelters for people experiencing homelessness

**DOI:** 10.1101/2022.01.14.22269074

**Authors:** Thomas D. Brothers, Malcolm Leaman, Matthew Bonn, Dan Lewer, Jacqueline Atkinson, John Fraser, Amy Gillis, Michael Gniewek, Leisha Hawker, Heather Hayman, Peter Jorna, David Martell, Tiffany O’Donnell, Helen Rivers-Bowerman, Leah Genge

**Author notes:** **Address correspondence to:** Thomas D. Brothers, MD. These authors listed in alphabetical order (Jacqueline Atkinson, John Fraser, Amy Gillis, Michael Gniewek, Leisha Hawker, Heather Hayman, Peter Jorna, David Martell, Tiffany O’Donnell).

## Abstract

**Background:** During a COVID-19 outbreak in the congregate shelter system in Halifax, Nova Scotia, Canada, a multidisciplinary health care team provided an emergency “safe supply” of pharmaceutical-grade medications and beverage-grade alcohol to facilitate isolation in COVID-19 hotel shelters for residents who are dependent on these substances. We aimed to evaluate (a) substances and dosages provided, and (b) effectiveness and safety of the program.

**Methods:** We retrospectively reviewed medical records of all COVID-19 isolation hotel shelter residents during May 2021. We extracted data on medication and alcohol dosages provided each day. The primary outcome was residents prematurely leaving isolation against public health orders. Adverse events included (a) overdose; (b) intoxication; and (c) diversion, selling, or sharing of medications or alcohol.

**Results:** Over 25 days, 77 isolation hotel residents were assessed (mean age 42 ± 14 years; 24% women). Sixty-two (81%) residents were provided medications, alcohol, or cigarettes. Seventeen residents (22%) received opioid agonist treatment medications (methadone, buprenorphine, or slow-release oral morphine) and 27 (35%) received hydromorphone tablets. Thirty-one (40%) residents received stimulant tablets with methylphenidate (27; 35%), dextroamphetamine (8; 10%), or lisdexamfetamine (2; 3%). Six residents (8%) received benzodiazepines. Forty-two (55%) residents received alcohol, including 41 (53%) with strong beer, three (3%) with wine, and one (1%) with hard liquor. Over 14 days in isolation, mean daily dosages increased of hydromorphone (45 ± 32 to 57 ± 42mg), methylphenidate (51 ± 28 to 77 ± 37mg), dextroamphetamine (33 ± 16 to 46 ± 13mg), and alcohol (12.3 ± 7.6 to 13.0 ± 6.9 standard drinks). Six residents (8%) left isolation prematurely, but four of those residents returned. Over 1,059 person-days in isolation, there were zero overdoses. Documented concerns regarding intoxication occurred six times (0.005 events/person-day) and medication diversion or sharing three times (0.003 events/person-day).

**Conclusions:** An emergency safe supply and managed alcohol program, paired with housing, was associated with low rates of adverse events and high rates of successful completion of the 14-day isolation period in COVID-19 isolation hotel shelters. This supports the effectiveness and safety of emergency safe supply prescribing and managed alcohol in this setting.

## BACKGROUND

The COVID-19 pandemic and associated public health restrictions have had a disproportionate impact on people who use drugs and/or alcohol (1–3). Disruptions to drug supply routes have led to an increasingly toxic and unpredictable drug supply, while physical distancing requirements cause more people to use drugs alone (where they cannot be resuscitated if they overdose) and have reduced capacity and operating hours at harm reduction and addiction treatment programs (4–6). People who are dependent on substances may be unable to follow public health directives to isolate if they have been exposed to COVID-19, due to withdrawal symptoms or compulsive use (1). For people who use drugs and/or alcohol and are also experiencing homelessness, staying in congregate shelters increases risks of COVID-19 infection; people in this situation would be unable to isolate unless given a private place to stay (7,8).

To facilitate physical distancing and decrease risks of COVID-19 infection, withdrawal, and overdose, Canadian clinicians developed rapid guidelines to provide a regular, safe supply of pharmaceutical-grade drugs and of beverage-grade alcohol to people who are dependent on these substances (9–12). The rationale for providing an alternative “safe supply” of substances to remove harms caused by reliance on the criminalized, unregulated, and poisonous drug market was first advanced by the Canadian Association of People who Used Drugs (CAPUD)(1,13,14) and developed clinically by Sereda and colleagues(15) and by Tyndall and colleagues(16), before the COVID-19 pandemic. Provisional prescribing of safe supply medications and managed alcohol to facilitate COVID-19 related physical distancing or isolation has also been termed “risk mitigation” or “pandemic prescribing” (1,9,17–19). The uptake of these prescribing guidelines on a population level is under evaluation (17,20), but the clinical safety and effectiveness of this approach for people in COVID-19 isolation has not been demonstrated.

In May 2021, there was a COVID-19 outbreak in the congregate shelter housing system in Halifax, Nova Scotia, Canada, and all residents in shelters experiencing COVID-19 outbreaks were moved to isolation hotel shelters for 14 days. A multidisciplinary health care team provided emergency, temporary safe supply medications and beverage-grade alcohol to facilitate isolation for residents who are dependent on these substances.

We aimed to describe the organization and delivery of an emergency, provisional safe supply drug and managed alcohol program during a COVID-19 outbreak in the congregate shelter system in Halifax, Nova Scotia, Canada. We evaluated safety of the program through the frequency of substance-related adverse events (including fatal and non-fatal overdose), and effectiveness through the rate of premature resident-initiated discharge from isolation against Public Health orders.

## METHODS

### Setting and data sources

This study comprises a retrospective case series of all COVID-19 isolation hotel shelter residents admitted during the Spring 2021 COVID-19 outbreak in the congregate shelter system in Halifax, Nova Scotia. This manuscript is reported in accordance with the Strengthening The Reporting of Observational studies in Epidemiology (STROBE) checklist (21).

People who stayed at shelters identified to have COVID-19 outbreaks were moved to isolation in hotels funded by the provincial government. At this stage in the pandemic, they were mandated to isolate for 14 days under authority of the Nova Scotia *Health Protection Act*. Isolation hotel shelters were in the city centre, several blocks away from residents’ usual congregate shelters. Residents of a given shelter typically stayed on the same hotel floor, with shelter staff continuing to support them there.

Data were extracted from the shared electronic medical record, including progress notes, electronic prescriptions, and messaging. Using structured chart review, each resident’s information was extracted in duplicate, once by a graduate student researcher (ML) and once by a clinician with experience prescribing these medications (TDB, MG, or AG). Discrepancies were resolved by TDB.

### Program description

Mobile Outreach Street Health (MOSH) organized a team of physicians and nurse practitioners with experience in addiction medicine and harm reduction, established a weekly clinical care coverage schedule, and provided access to a shared digital electronic medical record. MOSH was established in 2009 to provide outreach primary care to people experiencing homelessness and people who use drugs in Halifax; the organization has long-standing relationships with the city’s shelters and many of the residents. All residents being moved to isolation were referred to the harm reduction prescribing team for assessment. Nurses, nurse practitioners, and physicians performed intake assessments on substance use and health history; most assessments were done over the phone, but some were done in person.

Prescribers had access to province-wide pharmacy information system to confirm patient reports of prescribed medications, including opioid agonist treatment (OAT). Some patients were previously seen by MOSH or the associated North End Community Health Centre, and in this case had existing medical records the team could access.

Physicians and nurse practitioners prescribed pharmaceutical-grade substances generally following the BC Centre on Substance Use (BCCSU) Guidelines: Risk Mitigation in the Context of Dual Public Health Emergencies document (9), and beverage-grade alcohol according to MOSH managed alcohol program’s protocols. See Table 1 for a summary of prescribing guidance used by the MOSH team. Residents were aware that both the hotel-based private housing and the safe supply medications would only be provided for 14 days while they were isolating under Public Health orders.

**Table 1.** Summary of prescribing guidelines used in emergency safe supply drug and managed alcohol program in COVID isolation hotels in Halifax.

| Substance | Summary of prescribing guidance |
| --- | --- |
| Opoids | • Offer OAT to all patients with opioid use disorder.<br>• It is helpful to prescribe a long-acting opioid (e.g. slow-release oral morphine) in conjunction with a short-acting opioid for those not on OAT.<br>• Oral hydromorphone 8mg tablets, 1-3 tablets every hour as needed.<br>• Maximum daily dose of 14 tablets (112mg). |
| Stimulants | • Methylphenidate SR 20-40mg tablets once daily and/or methylphenidate IR 10-20mg tablets twice daily.<br>• Maximum daily dose of 100mg methylphenidate.<br>• Dextroamphetamine SR 10-20mg tablets twice daily and/or dextroamphetamine IR 10-20mg tablets twice or thrice daily.<br>• Maximum daily dose of 80-120mg dextroamphetamine. |
| Benzodiazepines | • If temporary maintenance is being prescribed, generally consider switching to a long-acting benzodiazepine (e.g. diazepam or clonazepam) and reduce dose by 50% to start and then titrate daily. |
| Alcohol | • Convert patient-reported alcohol consumption into "Canadian Standard Drinks".<br>• Most mouthwash estimated at 26% ABV, regular wine at 12% ABV, and fortified wine at 20%.<br>• Prescribe managed alcohol dose in number of cans of strong beer (6% ABV; 1.25 standard drinks per can) or red wine (12% ABV; 5.2 standard drinks per 750mL bottle). Limited hard liquor (40% ABV; 0.69 standard drinks per ounce) was also available on a case-by-case basis.<br>• Preference is to use beer, as it can be more easily spread throughout the day. |
| Tobacco | • Offer nicotine replacement therapy (i.e., patch, gum, lozenge, inhaler).<br>• Residents requiring tobacco would be delivered 1-2 packs of cigarettes daily by a local harm reduction organization outreach team. |

SR: sustained-release formulation. IR: immediate release formulation. ABV: Alcohol by volume.

The BCCSU guidelines were developed in British Columbia, which has a more potent and unpredictable illicit drug supply than Nova Scotia; as a result, prescribers did not know whether the recommended dosing ranges in the BCCSU guidelines would be required or appropriate. Compared to Nova Scotia, British Columbia has much higher rates of illicitly manufactured fentanyl, fentanyl analogues, novel benzodiazepines, and methamphetamine availability and use (22). People who use drugs in Nova Scotia most often use hydromorphone tablets (immediate release or extended release) and cocaine, though rates of illicitly manufactured fentanyl use are increasing (23–26). Largely due to these regional differences in the illicit drug supply, British Columbia experienced a rate of opioid poisoning deaths (39.4 per 100,000 people) eight times higher than Nova Scotia (4.9 per 100,000) from January to June, 2021 (26).

Prescribed medications could be taken orally, or crushed and injected or snorted; prescribers reviewed with residents that oral tablets were not designed to be crushed and injected, and provided guidance on safer use within a harm reduction framework. Resident preferences as to specific brands or formulations (e.g. those that might be more soluble in water to facilitate safer injecting) were followed as closely as possible. Liquid hydromorphone for injection use is not included in the BCCSU guidelines and was not considered here; this oversight has been criticized by people who use drugs because of the relatively increased harms associated with injecting oral tablets (27). Cannabis withdrawal is not mentioned in the BCCSU guidelines and the prescribing team initially underappreciated the importance of cannabis cravings and withdrawal symptoms (28), once other needs were met. While trying to facilitate funding for cannabis deliveries to the hotels, prescribers began to offer nabilone as an agonist replacement therapy to residents with cannabis withdrawal symptoms and residents began to order their own cannabis.

Medications were delivered daily by a local community pharmacist with experience with OAT and a harm reduction philosophy of care. Alcohol was delivered daily by the MOSH managed alcohol program outreach team or dispensed by shelter staff on site. For residents who reported intense binge drinking, alcohol dispensing would be divided into two times per day. Prescribers performed frequent phone follow-ups to adjust dosages, usually daily for the first three days and then as needed. MOSH nurses and/or prescribers would assess residents in person if needed. The team communicated via mobile secure messaging app and discussed challenging cases by phone and virtual video conferences. Mainline Needle Exchange, a local harm reduction outreach organization, provided all residents receiving safe supply medications with take-home naloxone kits, sterile drug preparation and injecting equipment, and support. No dedicated safe consumption space was created; instead, residents were encouraged to try “virtual spotting”(29) with friends or family or with the National Overdose Response Service (NORS) phone line (30), or otherwise to let shelter staff know they were going to be using so they could check in soon after.

There were no costs to residents at the COVID-19 isolation hotels. Medications were covered either through public drug insurance plans (for those who were enrolled) or by Nova Scotia Public Health (for those without insurance). Alcohol costs were initially covered by the MOSH managed alcohol program, and then through provincial government funding. Sterile injecting equipment and take-home naloxone kits are free to everyone in Nova Scotia, funded by the provincial government.

### Measures

#### Descriptive characteristics

We extracted data on resident demographic characteristics including age and gender. Race and Indigenous status were not routinely evaluated in the medical assessments and therefore were not available for extraction in the medical record. We extracted data on dosages of medications dispensed and calculated daily dosages and averages among patients receiving the medications. Alcohol was converted into Canadian standard drink units (17.05mL or 0.5765oz of pure ethanol).(31)

#### Primary outcome

The primary outcome was the frequency of residents leaving the isolation hotel shelter against public health orders before the mandatory 14 day isolation period was completed.

#### Adverse events

We extracted data on adverse events including documentation of (a) overdose; (b) intoxication; and (c) diversion, sharing, or selling of safe supply medications or alcohol.

Overdose was defined as fatal or non-fatal drug or alcohol poisoning that would require basic life support, administration of naloxone or oxygen, and/or transfer to the emergency department. Intoxication and diversion, sharing, or selling was documented in medical records as part of prescribers’ assessment and plan to continue or change dosages of medications and alcohol, based on prescribers’ clinical impression (usually by telephone), by resident report, or by *ad hoc* descriptions by shelter support staff, the pharmacist, or the managed alcohol program outreach team.

### Analysis

We used Microsoft Excel for data management and to calculate summary statistics and R 3.6.3 for data visualizations. We described individual trajectories by creating separate plots for each resident’s daily dosages of opioids, stimulants, and alcohol. To compare different substances on the same visual scale, we transformed individual’s daily dosages into a percentage of the maximum daily dosage of that substance received across the whole sample; for example, the maximum daily hydromorphone dosage across all residents was 158mg, so an individual resident receiving 48mg of hydromorphone in a day would have a percentage value of 16mg ÷ 158mg x 100% = 30% for that day.

## RESULTS

### Participants

Over 25 days, 77 residents were admitted to COVID-19 isolation hotel shelters and referred to the medical team (Table 2). In total, there were 1,059 person-days in isolation after medical assessment. Most participants were men, and average age differed by gender. Mean age for men was 46 ± 14 years, and for women was 30 ± 10 years. After intake assessment, 15 residents (19%) were determined to have no concerns about substance withdrawal or dependence while in isolation and were given no medications, alcohol, or cigarettes. Sixty-two residents (81%) were provided medications, alcohol, or cigarettes, summarized by day of isolation in Figure 1.

**Table 2.** Descriptive characteristics of the sample of residents in COVID-19 isolation.

|  |  |
| --- | --- |
| Sample size (n) | 77 |
| Age, years (mean ± standard deviation) | 42 ± 14 |
| Gender, women (%) | 19 (25%) |
| Residents provided opioid agonist treatment (n) |  |
| Any opioid agonist treatment | 17 (22%) |
| Methadone | 7 (9%) |
| Buprenorphine-naloxone | 1 (1%) |
| Slow-release oral morphine | 10 (13%) |
| Residents provided hydromorphone (n) | 27 (35%) |
| Residents provided benzodiazepines (n) |  |
| Any benzodiazepine | 6 (8%) |
| Clonazepam | 5 (6%) |
| Lorazepam | 1 (1%) |
| Residents provided stimulants (n) |  |
| Any stimulant | 31 (40%) |
| Methylphenidate | 27 (35%) |
| Dextroamphetamine | 8 (10%) |
| Lisdexamfetamine | 2 (3%) |
| Residents provided alcohol (n) |  |
| Any alcohol | 42 (55%) |
| Strong beer (6% ABV) | 41 (53%) |
| Wine (12% ABV) | 3 (4%) |
| Liquor (40% ABV) | 1 (1%) |
| Residents provided nicotine replacement therapy (n) | 2 (3%) |
| Residents provided cigarettes (n) | 64 (83%) |
| Residents provided nabilone (n) | 14 (18%) |

ABV: alcohol by volume.

**Figure 1.**
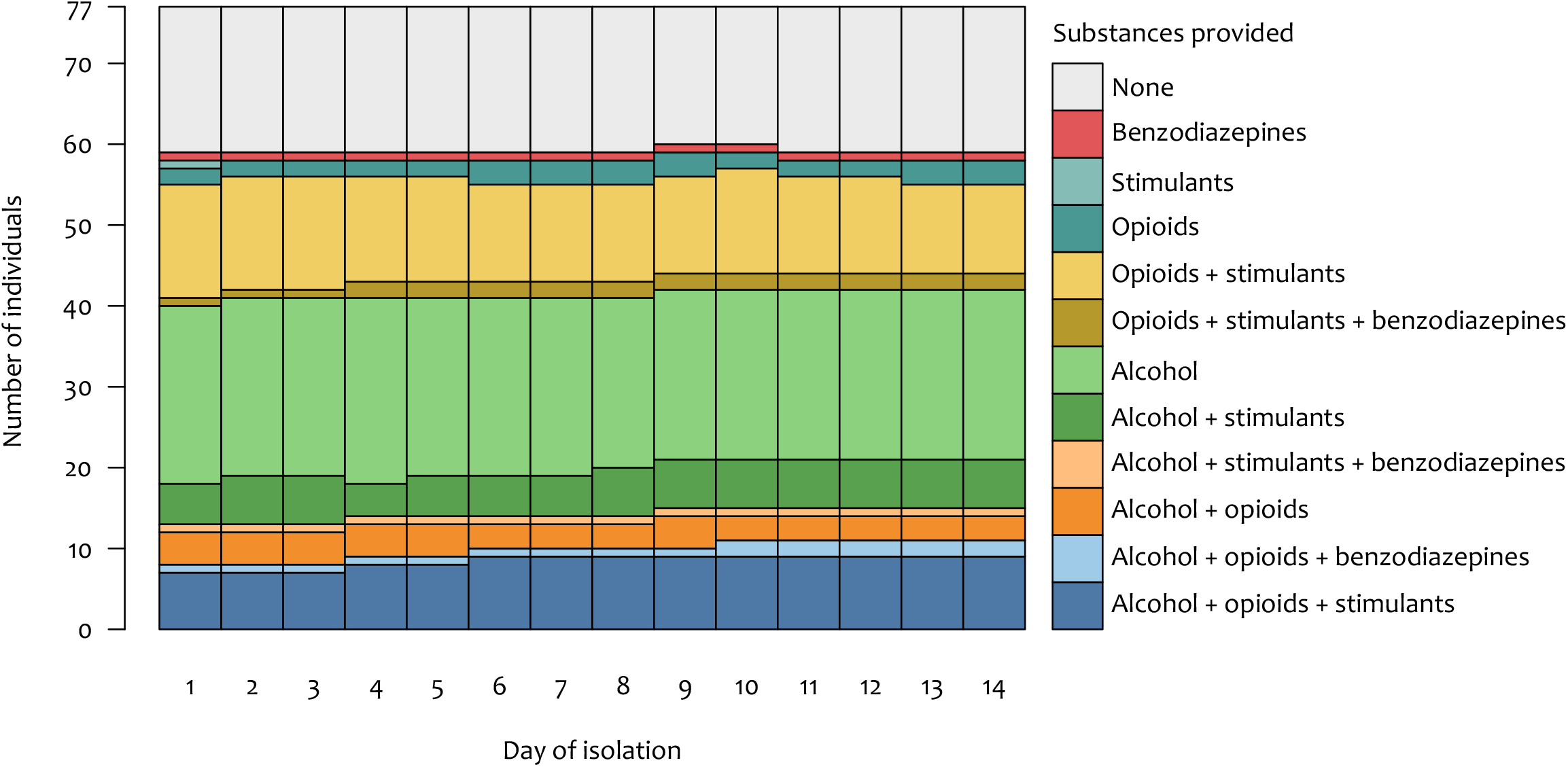
Number of COVID-19 isolation hotel shelter residents receiving each category of safe supply medications or managed alcohol during 14 days of isolation. Benzodiazepines include clonazepam and lorazepam. Stimulants include methylphenidate, dextroamphetamine, and lisdexamfetamine. Opioids include opioid agonist treatment medications (methadone, buprenorphine, or slow-release morphine) and hydromorphone. Alcohol includes strong beer, wine, or hard liquor.

Cigarettes were the most commonly provided substance (64 residents; 83% of total sample), followed by alcohol (42 residents; 55% of total sample), and hydromorphone tablets (27 residents; 35% of total sample). Seventeen residents (22%) received any OAT, including eight who initiated OAT medications in the isolation hotel shelters. All eight of these residents initiated SROM, and no residents initiated methadone or buprenorphine-naloxone. Twelve residents received both OAT and hydromorphone tablets on the same day (71% of residents receiving OAT); four of these residents were already on OAT before isolation. Two residents accepted offers of nicotine replacement therapy, including one resident who also had cigarettes delivered.

### Safe supply medication and managed alcohol dosages

Among the 27 residents receiving hydromorphone, average dosages increased over residents’ time in isolation from day one (mean 45mg ± 32mg; median 32mg; range 16 - 158mg daily) to day 14 (mean 57mg ± 42mg; median 48mg; range 16 - 158mg daily) (Supplementary Figure S1). Three (12%) of these 27 residents were prescribed hydromorphone dosages above the BCCSU guideline suggested upper limit of 112mg daily (14 × 8mg tablets). Individual daily dosage trajectories for hydromorphone and OAT are visualized in Figure 2, plotted as percentages of the maximum daily dosage of each medication across the whole sample. The maximum daily dosage for methadone was 195mg, for buprenorphine was 12mg, for SROM was 800mg; and for hydromorphone was 158mg.

**Figure 2.**
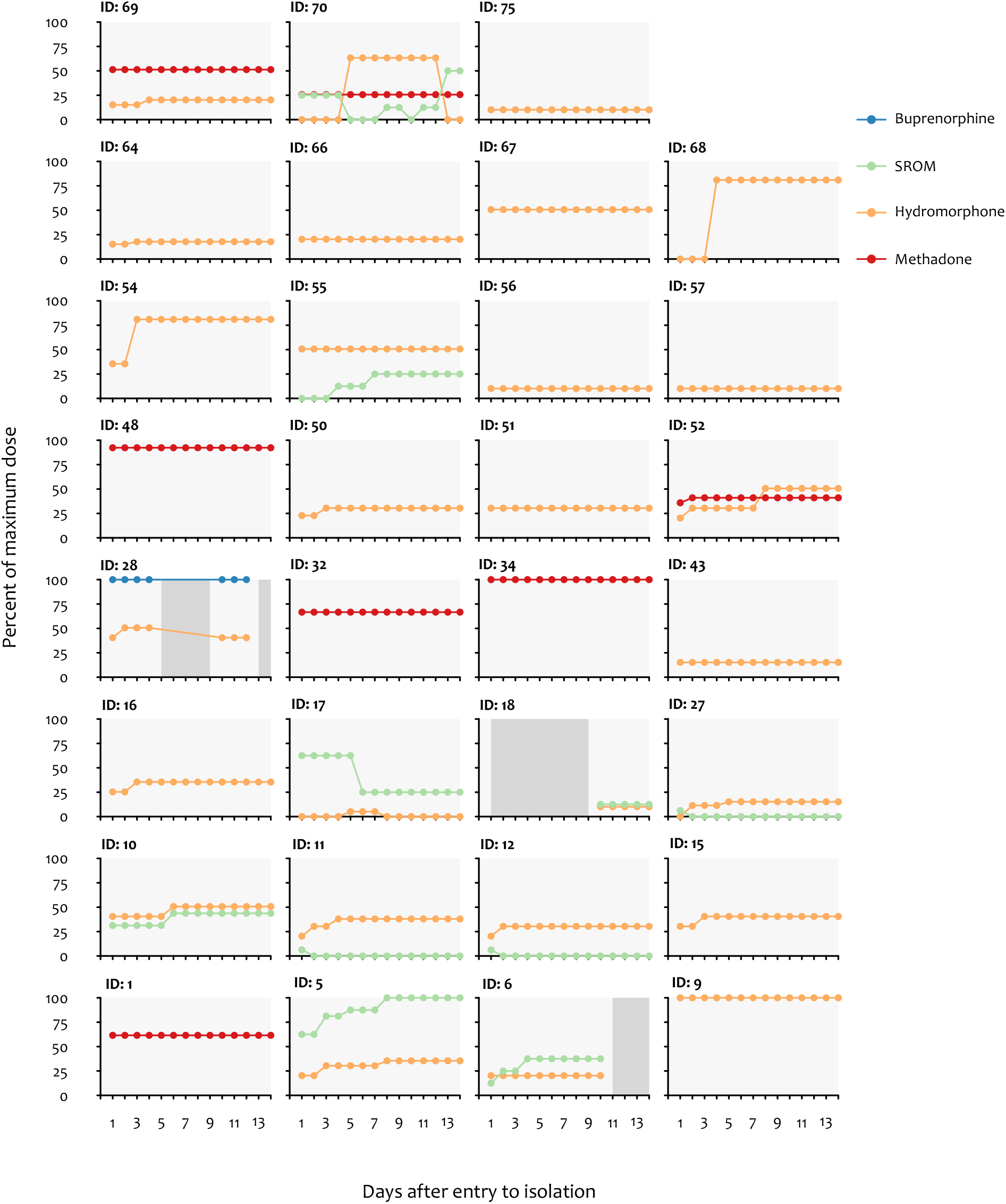
COVID-19 isolation hotel shelter residents’ daily dosage trajectories for safe supply hydromorphone and opioid agonist treatment medications. Individual daily medication dosages are plotted as percentages of the maximum daily dosages of each substance across the whole sample (i.e., methadone 195mg; buprenorphine 12mg; SROM 800mg; hydromorphone 158mg). Dark grey boxes represent days where substances could not be provided, either because resident was away from isolation or because of delayed medical assessment. SROM: slow-release oral morphine.

Among residents receiving stimulants, average dosages also increased over time (Supplementary Figure S2). Methylphenidate daily dosages increased from day one (mean 51mg ± 28mg; median 40mg; range 10 - 107mg) to day 14 (mean 77mg ± 37mg; median 80mg; range 15 - 160mg). Dextroamphetamine daily dosages increased from day one (mean 33mg ± 16mg; median 30mg; range 20 – 60mg) to day 14 (mean 46mg ± 13mg; median 40mg; range 30 – 60mg). Four (15%) of 27 residents receiving methylphenidate were prescribed doses above the BCCSU guideline suggested upper limit of 100mg daily. Of eight residents receiving dextroamphetamine, one (13%) required dosages above the guideline suggested upper limit of 120mg daily. Individual daily dosage trajectories for stimulant medications are visualized in Figure 3, plotted as percentages of the maximum daily dosage of each medication across the whole sample (methylphenidate 160mg, dextroamphetamine 80mg, and lisdexamfetamine 60mg).

**Figure 3.**
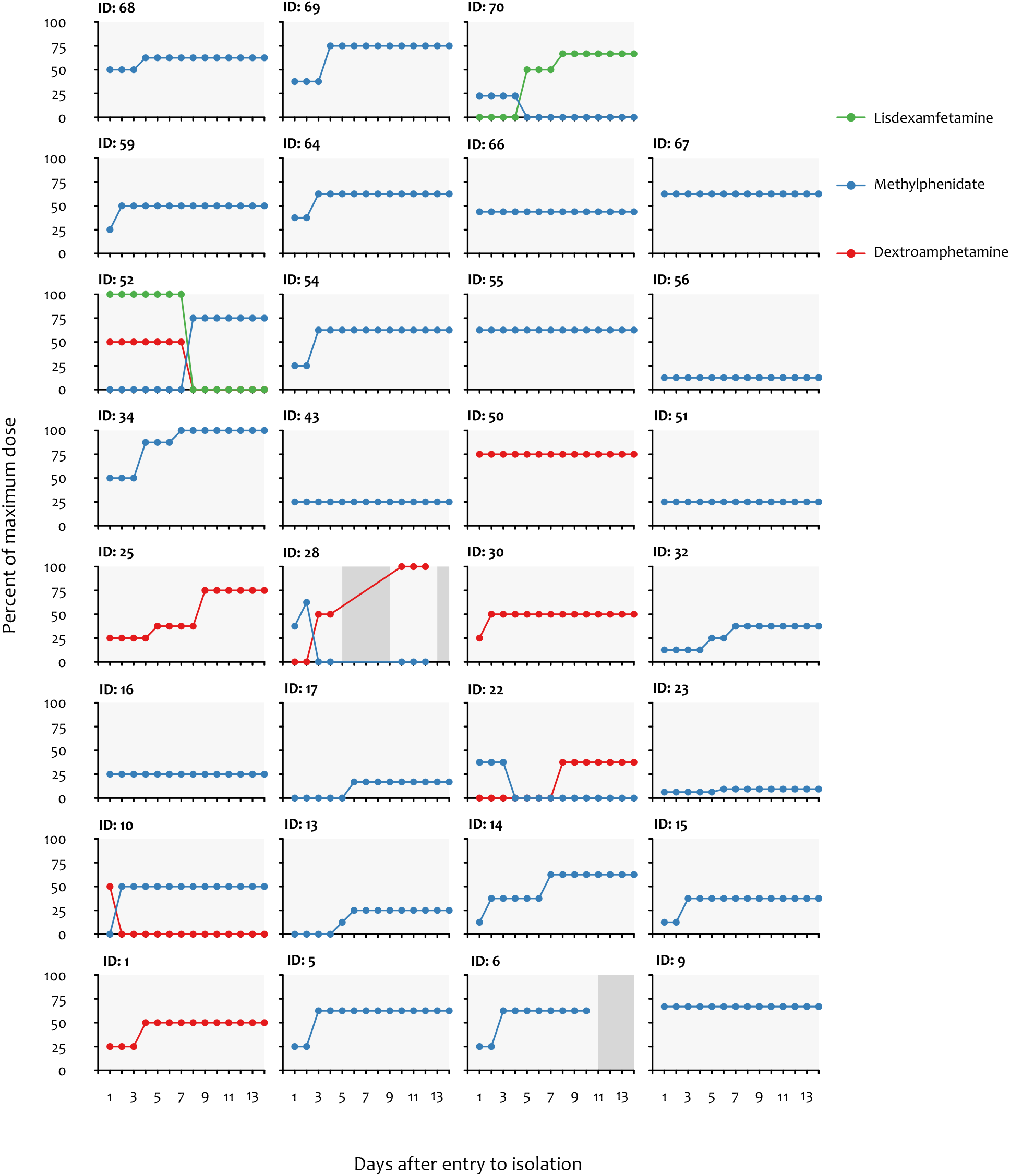
COVID-19 isolation hotel shelter residents’ daily dosage trajectories for safe supply stimulant medications. Individual daily medication dosages are plotted as percentages of the maximum daily dosages of each substance across the whole sample (i.e., methylphenidate 160mg; dextroamphetamine 80mg; lisdexamfetamine 60mg). Dark grey boxes represent days where substances could not be provided, either because resident was away from isolation or because of delayed medical assessment.

Average daily alcohol dosages increased slightly over time from day one (mean 12.3 ± 7.6 standard drinks; median 11.25 standard drinks; range 1.25 - 33.75 standard drinks) to day 14 (mean 13.0 ± 6.9 standard drinks; median 13.1 standard drinks; range 1.25 - 30.75 standard drinks) (Supplementary Figure S3). Individual daily dosage trajectories for alcohol are visualized in Figure 4, plotted as percentages of the maximum daily dosage of alcohol across the whole sample (37.5 standard drinks).

**Figure 4.**
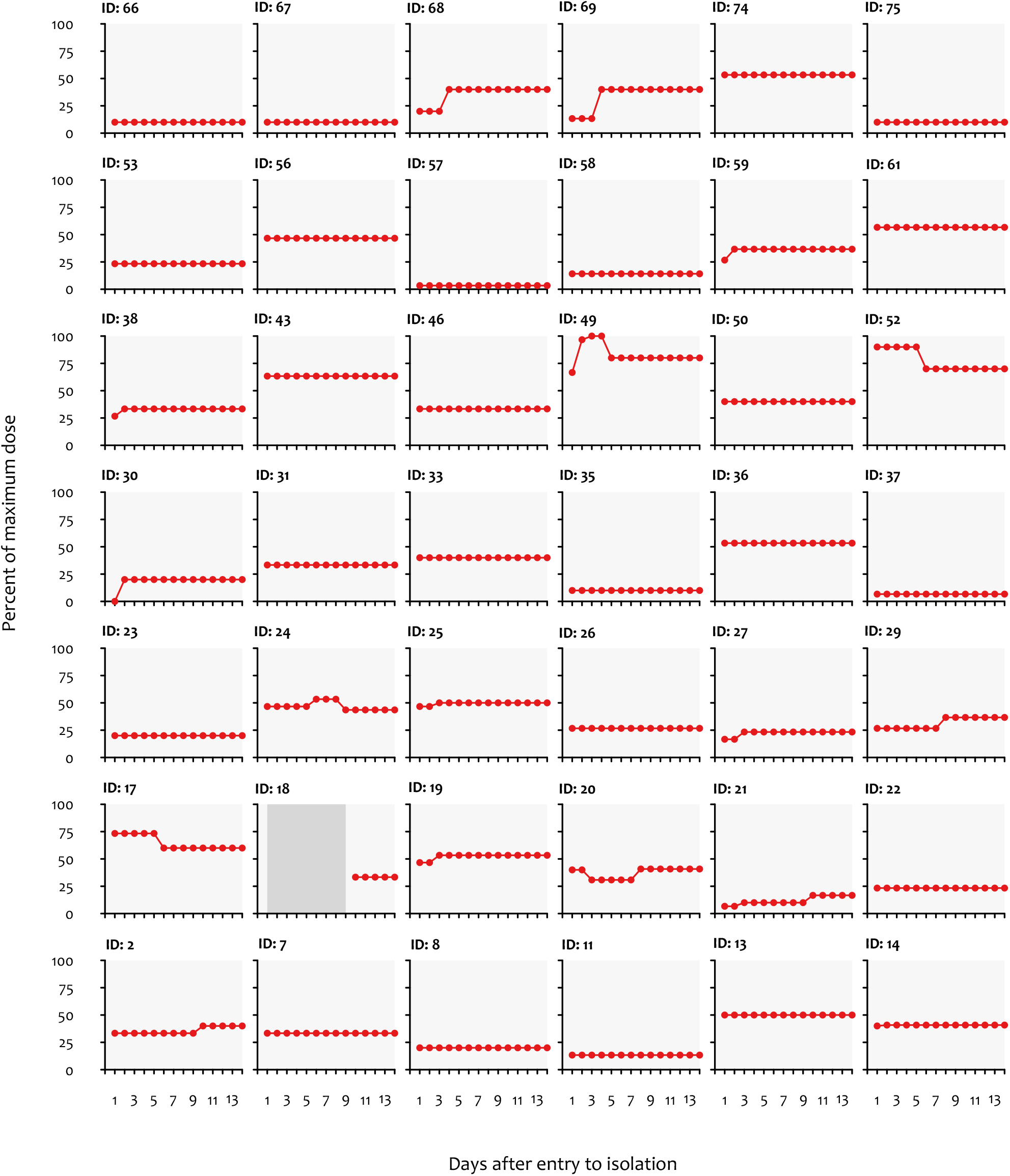
COVID-19 isolation hotel shelter residents’ daily dosage trajectories for managed alcohol. Individual daily standard drink dosages are plotted as percentages of the maximum daily dosage of alcohol across the whole sample (i.e., 37.5 standard drinks). Dark grey boxes represent days where substances could not be provided, either because resident was away from isolation or because of delayed medical assessment.

Benzodiazepine dosages were relatively stable. Clonazepam increased slightly from day one (mean 1.67 ± 1.15; median 1mg; range 1 – 3mg) to day 14 (mean 2.00 ± 1.41mg; median 1mg; range 1 - 4mg) and the only lorazepam daily dosage was stable at 1mg. Nabilone dosages increased from mean 2mg ± 0mg on day one to mean 2.79mg ± 1.25mg on day 14, while an unknown number of residents had cannabis delivered to the isolation hotel shelters.

### Primary outcome

Among the 77 isolation hotel residents, six (8%) left against public health orders. Four of these six soon returned and remained in isolation, resulting in two (3%) persistent premature discharges from isolation.

### Adverse events

Over 1,059 person-days in isolation, there were zero overdoses in the isolation hotel shelters. Concerns regarding intoxication were documented six times (0.005 events per person-day); four of these residents with documented intoxication were provided alcohol and four were provided opioids (three with OAT plus hydromorphone, and one with hydromorphone only). Concerns regarding diversion, sharing, or selling of medications was documented three times (0.003 events per person-day), including among two residents who also had documented intoxication. All three of these residents were provided multiple substances, including opioids, stimulants, and alcohol.

## DISCUSSION

Among residents of a COVID-19 isolation hotel shelter for people experiencing homelessness, we found that an emergency, provisional safe supply program (i.e. prescribing pharmaceutical-grade medications and beverage-grade alcohol) was associated with low rates of adverse events and high rates of successful completion of the 14-day isolation period. No shelter residents experienced an overdose during their stay. We identified medication dosage ranges that generally fell within those recommended in “risk mitigation” prescribing guidelines, which were urgently produced in response to evolving risks of COVID-19. This supports the safety and effectiveness of this approach in this setting.

The safe supply drug and alcohol prescribing practices described in this evaluation are a recent development. While the relative safety of medications and alcohol dispensed for unwitnessed consumption has not been previously well-described in the literature, the practice is an extension of the evidence from witnessed consumption settings (14,15,18,25,32,33). Witnessed injectable OAT (iOAT) with liquid hydromorphone or diacetylmorphine (Heroin) has a robust evidence-based and has been incorporated into Canadian clinical practice guidelines for opioid use disorder (34,35). Qualitative studies have evaluated the benefits of witnessed hydromorphone tablet consumption, which is more flexible and less resource-intensive than witnessed iOAT (36,37). A recent study from Ottawa, Canada, describes positive outcomes for people with severe opioid use disorder who are provided hydromorphone iOAT along with supported housing (38). Benefits of managed alcohol programs are also clearly established for people with severe alcohol use disorder, and particularly people who drink non-beverage alcohol (39–41). Some existing managed alcohol programs include daily and/or unwitnessed ingestion (42).

The dosing strategy informed by the BCCSU guidelines were appropriate for most patients in this setting (Halifax, Nova Scotia) where the illicit drug supply is comprised primarily of pharmaceutical hydromorphone and of cocaine, with relatively little fentanyl and methamphetamine availability in the community (23,24,43,44). In other settings, dosages may need to be higher than those recommended in these guidelines or different medications may be for effective. For example, a recent survey of people who use drugs in British Columbia, Canada, showed that in that province most would prefer heroin or fentanyl safe supply over prescription opioids like hydromorphone (45). For the emergency safe supply program in Halifax described in our study, many residents were able to report their usual daily use of non-prescribed hydromorphone tablets which could be matched with the safe supply prescription. While the mean dosages of hydromorphone, methylphenidate, and dextroamphetamine increased over residents’ 14 days in isolation, many patients stayed at the same dose throughout. As there were no overdoses and very few premature discharges from isolation, this suggests that residents knew how much medication they would need and were willing to work with the prescriber if started too low. While these medications were not offered as substance use disorder treatment, the options available to patients (in terms of medications, dosages, and brands or formulations) to help facilitate goals of successful 14 day isolation represented elements of shared decision-making and patient-centered care (25,46,47). It is notable that with the broad selection of options available to avoid reliance on the criminalized drug supply, no residents chose to start methadone or buprenorphine OAT. This differs from other settings like acute care hospitals, where patients with medical complications of opioid use disorder may not initially be treatment-seeking, but are often motivated to engage in OAT when offered (24,25,48). Prior research in the hospital setting has shown that offering SROM in addition to methadone and buprenorphine may increase treatment uptake (25).

Descriptions of harm reduction practices in COVID-19 isolation shelters have been reported from Toronto(49) and Hamilton(50,51), Canada; Boston(49,52) and San Francisco(53,54), USA; Lisbon, Portugal (55); and Tshwane, South Africa (56). The other Canadian harm reduction programs are most like the one described in our study. The Toronto program supported isolation shelter residents with an emergency managed alcohol program, safe supply hydromorphone prescribing, opioid agonist treatment, take-home naloxone kits, sterile injecting equipment, and telephone or in-person check-ins; specific medication and alcohol dosages and frequencies are not reported. They established a supervised consumption site for witnessed injections, after applying for federal approval. The Toronto program reported 4 suspected overdose deaths among 1700 admissions (0.2%), which were all unwitnessed (49). The Hamilton program supported residents isolating at a men’s congregate shelter with sterile injecting equipment, take-home naloxone, a flexible OAT delivery model, and hydromorphone safe supply prescribing (51). A community organization set up a supervised consumption space within the shelter where residents could consume their prescribed hydromorphone. The Hamilton program reported no fatal overdoses and three non-fatal overdoses during the month-long intervention (all of which occurred outside the safe consumption site), compared to 20 non-fatal overdoses in the month before the isolation period (51). The Hamilton program description did not mention managed alcohol, and neither report includes the frequency of residents leaving isolation prematurely.

In Boston, OAT was offered but safe supply prescriptions and alcohol were not (49,52). Naloxone and sterile syringes were distributed at discharge from the isolation shelters, but not provided to residents during their stay. In San Francisco, prescribers offered OAT (with buprenorphine or methadone), medical cannabis, nicotine replacement therapy, and managed alcohol (53). Residents were provided with sterile injecting equipment and naloxone, and $20 gift cards for completing their stay. Opioids, stimulants, and benzodiazepines were not offered. Some San Francisco shelter programs limited managed alcohol to a maximum dosage of 10 standard drinks per day (54,54), which was below the mean and median dosages for the residents in Halifax in our study. Nineteen percent of San Francisco residents left isolation shelters prematurely (53), which was higher than the 3-8% in Halifax. In Lisbon, harm reduction organizations provided support, sterile injecting equipment, naloxone, and a mobile drug consumption room to shelter residents. Residents had access to benzodiazepines for alcohol withdrawal management, but no safe supply or managed alcohol program was provided (55). In Tshwane, prescribers provided methadone in the stadium-based emergency shelter. The local needle exchange program was asked not to deliver sterile injecting equipment, as “municipal and national police were actively confiscating needles and the city regarded the concurrent provision of needle and syringe services and [opioid substitution treatment] as a form of mixed messaging” (56).

The decision to revoke hotel-based private housing and safe supply medications after 14 days, despite the apparent benefits to individual residents and despite the ongoing COVID-19 pandemic, raises challenging ethical issues (57–59) and prevents evaluation of the potential long-term impact of these housing and safe supply interventions. These decisions were made by government and public health officials independent of the prescribers and study investigators.

Our study has important limitations. First, as the decision was made to offer all shelter residents this program for drug and alcohol withdrawal management, there is no control group of residents without this program to compare rates of adverse events or resident-initiated premature discharge from the isolation shelters against public advice. Nevertheless, the rate of premature discharge was lower here than reported in San Francisco, and our findings here of relatively safety are reassuring. Second, as our study relied on retrospective evaluation of medical records, we may be missing data on events (including medication diversion, sharing, or selling) that were not disclosed to shelter staff. The program described here did not have a systemic approach to surveillance or of gathering information on diversion, sharing, or selling from shelter staff. Other study designs, including qualitative interviews, could be used to get a better sense of the scale of medication diversion, sharing, and selling, that was not reported back to the medical team. Third, as our study occurred in a city with relatively little fentanyl and crystal methamphetamine use, the dosing ranges here may not be sufficient in populations with higher drug tolerance and this may limit generalizability.

## Conclusion

We found that an emergency, provisional safe supply program providing pharmaceutical-grade medications and beverage-grade alcohol in COVID-19 isolation hotel shelters was associated with low rates of adverse events and of high rates of successful completion of the mandatory 14-day isolation stay. This suggests this approach is safe and effective in this setting.

## Data Availability

All data produced in the present work are contained in the manuscript

## Abbreviations

ABV: Alcohol by volume
BCCSU: British Columbia Centre on Substance Use
MOSH: Mobile Outreach Street Health
NORS: National Overdose Response Service
OAT: Opioid agonist treatment
SROM: Slow-release oral morphine

## Declarations

### Ethics approval and consent to participate

Requirements for full ethics review and individual participant consent were waived by the Nova Scotia Health Research Ethics Board, who determined this project to be quality assessment (REB FILE #: 1027156).

### Consent for publication

Not applicable.

### Availability of data and materials

All data generated or analysed during this study are included in this published article.

### Author contribution

TDB contributed to conceptualization, developed and piloted the data extraction form, performed structured data extraction, validated data extraction, data curation, data analysis, data interpretation, co-wrote the first draft, and project supervision. ML developed and piloted the data extraction form, performed structured data extraction, data curation, and data analysis, and co-wrote the first draft. DL contributed to data analysis and interpretation. AG and MG contributed to conceptualization, data extraction, and interpretation. HR-B contributed to conceptualization, data entry, data curation, data interpretation, and project administration. MB, JA, JF, LH, HH, SH, PJ, DM, TO, and LG contributed to conceptualization and data interpretation. All authors provided critical intellectual input, contributed to revised drafts, and approved the final draft for submission.

## Acknowledgements

We acknowledge the essential and compassionate contributions of the wider MOSH team and shelter staff to the success of the isolation hotels.

We thank Dr. Claire Bodkin (McMaster University) and Dr. Ashish Thakrar (University of Pennsylvania) for helpful comments on earlier drafts.

