## Supplement for "Evaluation of an emergency safe supply drugs and managed alcohol program in COVID-19 isolation hotel shelters for people experiencing homelessness"

*These authors listed in alphabetical order

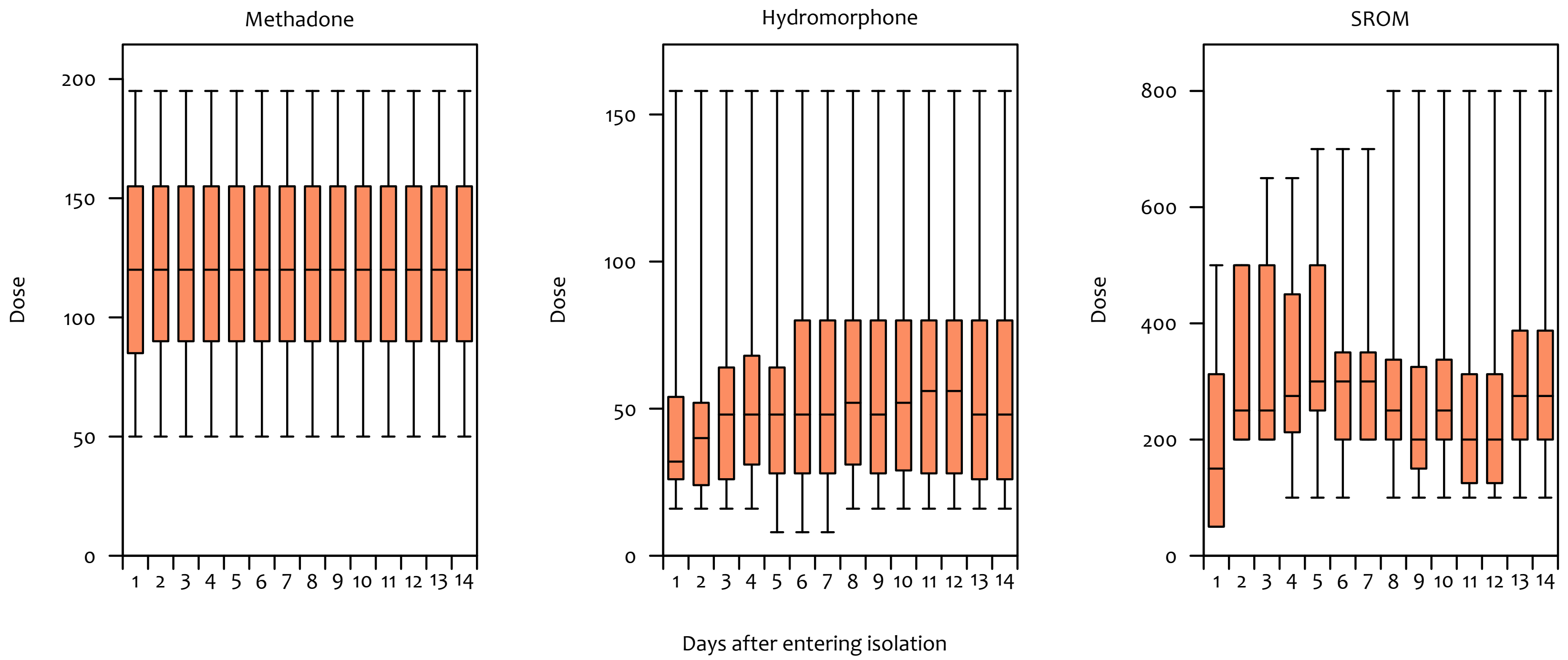


**Supplementary Figure S1. Boxplot summary of daily dosages of methadone, hydromorphone, and slow-release oral morphine (SROM) received by COVID-19 isolation hotel shelter residents.** Doses in milligrams.

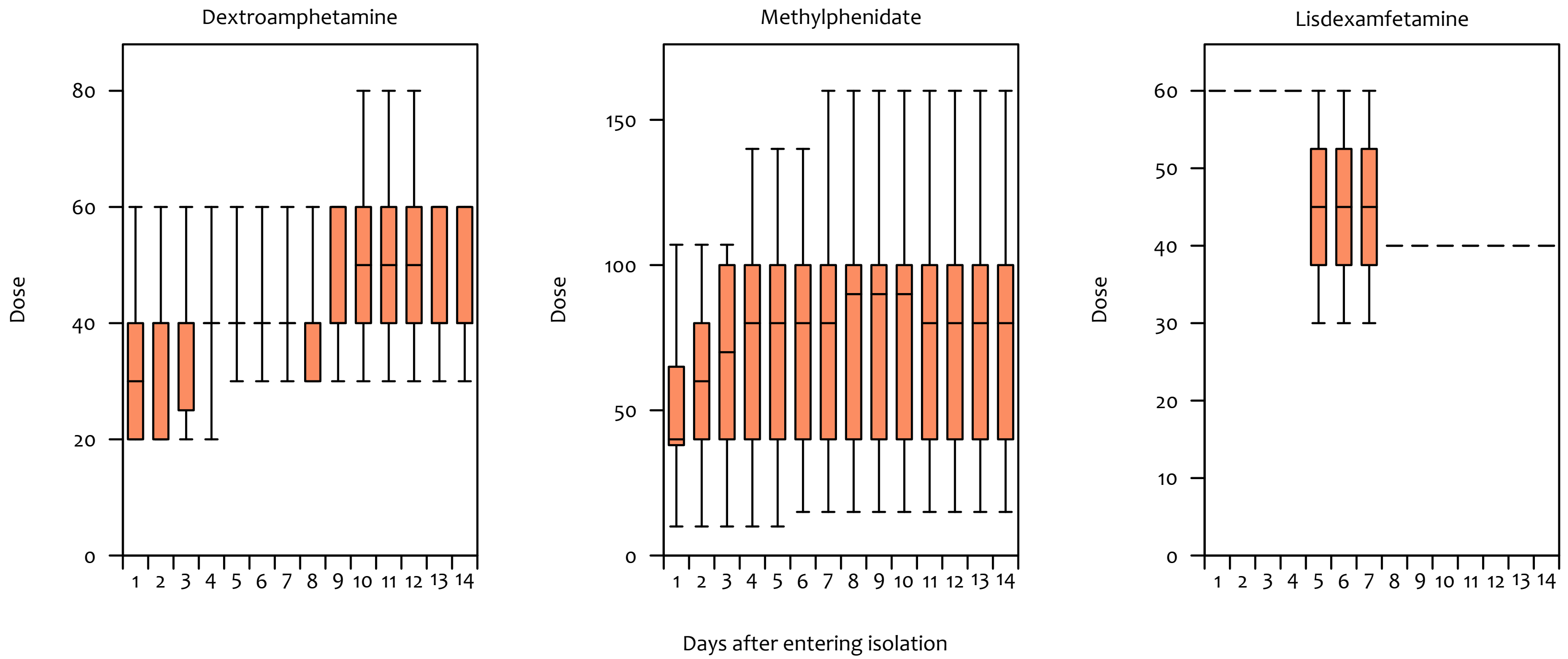


**Supplementary Figure S2. Boxplot summary of daily dosages of safe supply stimulant medications received by COVID-19 isolation hotel shelter residents.** Doses in milligrams.

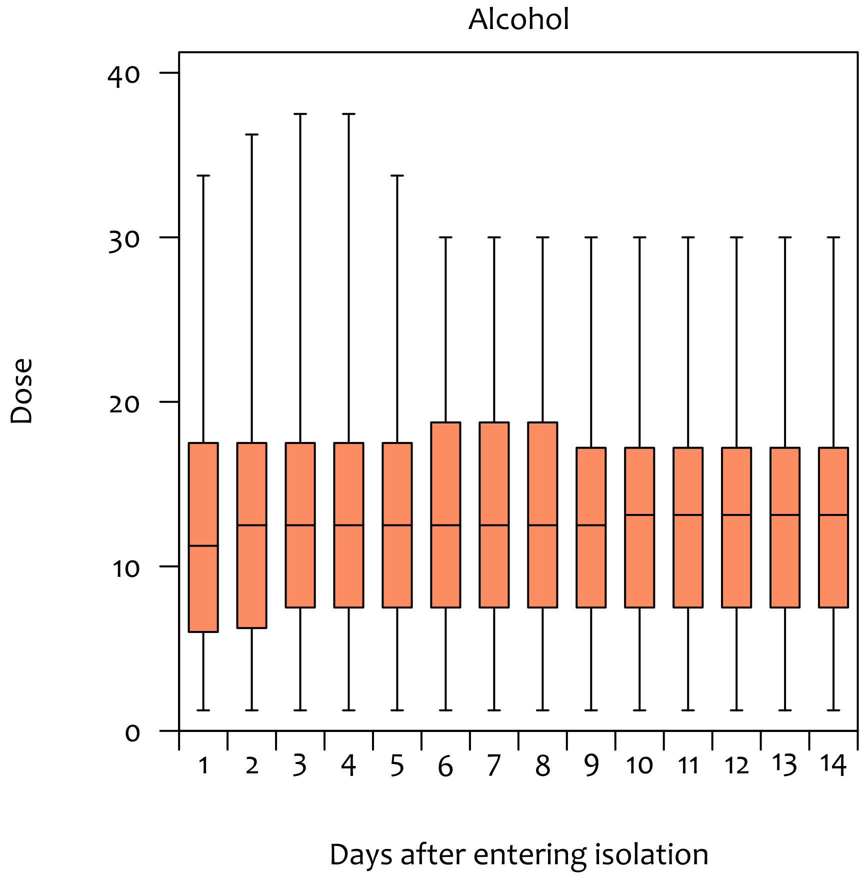


**Supplementary Figure S3. Boxplot summary of daily dosages of alcohol received by COVID-19 isolation hotel shelter residents.** Doses in standard drinks.
